# Integrated packages of non-pharmaceutical interventions increased public health response efficiency against COVID-19 during the first European wave: evidence from 32 European countries

**DOI:** 10.1101/2020.08.17.20174821

**Authors:** Andres Garchitorena, Hugo Gruson, Bernard Cazelles, Tommi Karki, Bertrand Sudre, Benjamin Roche

**Affiliations:** MIVEGEC, Univ. Montpellier, CNRS, IRD, Montpellier, France; IRD, Sorbonne University, UMMISCO, F-93143, Bondy, France; IBENS, CNRS UMR 8197, École Normale Supérieure, Paris, France; European Centre for Disease Control, Stockholm, Sweden, France; Departamento de Etología, Fauna Silvestre y Animales de Laboratorio, Facultad de Medicina Veterinaria y Zootecnia, Universidad Nacional Autónoma de México (UNAM), Ciudad de México, México

## Abstract

**Introduction:** Since the emergence of SARS-CoV-2, governments have implemented a combination of public health responses based on non-pharmaceutical interventions (NPIs), with significant social and economic consequences. Quantifying the efficiency of different NPIs implemented by European countries to overcome the first epidemic wave could inform preparedness for forthcoming waves.

**Methods:** We used a dataset compiled by the European Centre for Disease Control (ECDC) on daily COVID-19 incidence, mortality and NPI implementation in 32 European countries. We adapted a capture-recapture method to limit non-reporting bias in incidence data, which we fitted to an age-structured mathematical model coupled with Monte Carlo Markov Chain to quantify the efficiency of 258 public health responses (PHR, a combination of several NPIs) in reducing SARS-Cov-2 transmission rates. From these PHR efficiencies, we used time series analyses to isolate the effect of 13 NPIs at different levels of implementation (fully implemented vs. partially relaxed).

**Results:** Public health responses implemented in Europe led to a median decrease in viral transmission of 71%, enough to suppress the epidemic. PHR efficiency was positively associated with the number of NPIs implemented simultaneously. The largest effect among NPIs was observed for stay at home orders targeted at risk groups (β=0.24, 95%CI 0.16-0.32) and teleworking (β=0.23, 95%CI 0.15-0.31), followed by enforced stay at home orders for the general population, closure of non-essential businesses and services, bans on gatherings of 50 individuals or more, and closure of universities. Partial relaxation of most NPIs resulted in lower than average or non-significant changes in response efficiency.

**Conclusion:** This large-scale estimation of NPI and PHR efficiency against SARS-COV-2 transmission in Europe suggests that a combination of NPIs targeting different population groups should be favored to control future epidemic waves.

## Introduction

Since its emergence in China in December 2019, the SARS-COV-2 pandemic has affected almost every country in the World [1]. With more than 35 million cases reported and over one million deaths in the first 9 months of the pandemic[2], this virus will leave a lasting imprint in human history. To control national epidemic, many governments and national public health authorities have implemented national public health responses (PHR) that combine several non-pharmaceutical interventions simultaneously (NPIs). To ultimately achieve either a suppression or a mitigation of the epidemic, NPIs aim to reduce transmission by (i) lowering contact rates in the general population or specific groups through the curtailment of contact(s) with infectious individuals, and (ii) reduce the infectiousness of contacts. The NPIs implemented by most countries involve the closure of schools and universities, banning gatherings of various sizes, issuing stay at home recommendations or orders, closing businesses (e.g. hotels, restaurants) or services (e.g. public transportation, recreational activities) and the use of individual protective equipment (e.g., wearing masks), with different levels of enforcement [3]. Given the huge economic and social costs of these interventions, understanding their efficiency at reducing transmission can be key to better control future epidemic waves, allowing to timely prioritize association of NPIs.

Previously, mathematical models of COVID-19 suggested that lockdowns are the most efficient NPIs to reduce transmission [4] and may even be necessary to achieve epidemic suppression [5]. Using information on mobility patterns, other modelling studies predict that integrated responses that involve packages of several NPIs could have the strongest effects at reducing transmission, and coordinated public health responses across countries could delay subsequent epidemic waves [7,8]. Although there is large variability and uncertainty around the effect of individual NPIs within and across studies [5–12], an empirical analysis of local, regional and national implementation of NPIs in 6 countries in the first three months of the pandemic confirmed that packages of NPIs may achieve the largest reductions in SARS-COV-2 transmission [13]. Importantly, the empirical evidence currently available is limited by the small temporal and spatial representability of these initial studies.

The unprecedented range of public health responses implemented across European countries, in addition to variations in their combination, timing and level of NPI implementation provides an opportunity to perform a more robust quantification of their efficiency at reducing SARS-COV-2 transmission [4]. Using a dataset compiled by the European Centre for Disease Prevention and Control (ECDC) on country response measures to COVID-19 [14] paired with epidemiological incidence data [15], the goal of this study was to understand the efficiency of NPIs and public health responses (PHRs, a combination of NPIs) implemented in 32 European countries between February and September 2020.

## Materials and methods

### Data sources

We obtained data on COVID-19 incidence and NPI implementation for 32 European countries from February 1^st^ to September 16^th^, 2020. The data on COVID-19 incidence were retrieved from the ECDC epidemic intelligence database on worldwide data on COVID-19 [15]. It contains, for each country since the beginning of the epidemic, the total number of COVID-19 cases and deaths per day reported by each country. We also obtained information about NPI implementation over time as part of larger PHRs in each European country through the ECDC database on country response measures to COVID-19 [16]. It contains, for the same time period, the dates of beginning and end of PHR implementation in each country, comprising 13 individual NPIs (Table 1) and two levels of implementation for each NPI (fully implemented vs. partial relaxation of measures). This database is regularly updated by the ECDC using authoritative sources from national authorities and international institutions [14]. The ECDC used a template to update the data on NPIS every 15 days during a four days round where each national responsible review the recent NPIS based on the Ministry of Health website, other response measures database (<u>ACAPS</u>) and Health System Response Monitor (HSRM) from the European Observatory on Health Systems and Policies, which developed and maintains the platform, with support from the WHO Regional Office for Europe and the European Commission. Once all the information is triangulated, the data are consolidated and verified (database level). The information has been integrated for numerous rapid assessment and reports which are reviewed by the EU/EEA countries and European Commission.

**Table 1.** Description of Non Pharmaceutical Interventions (NPI) assessed

| **NPI Description** | **Abbreviation** |
| --- | --- |
| **Stay at Home Orders** |  |
| Stay-at-home orders for the general population (these are enforced and also referred to as ‘lockdown’) | StayHomeOrder |
| Stay-at-home recommendations for the general population (which are voluntary or not enforced) | StayHomeGen |
| Stay-at-home recommendations for risk groups or vulnerable populations (such as the elderly, people with underlying health conditions, physically disabled people, etc.) | StayHomeRiskG |
| **School/University Closures** |  |
| Closure of educational institutions: daycare or nursery | ClosDaycare |
| Closure of educational institutions: primary schools. | ClosPrim |
| Closure of educational institutions: secondary schools. | ClosSec |
| Closure of educational institutions: higher education. | ClosHigh |
| **Bans to Mass Gatherings** |  |
| Interventions are in place to limit mass/public gatherings (limit of 1000 or less individuals allowed). | MassGatherAll |
| Interventions are in place to limit mass/public gatherings (limit of 50 or less individuals allowed). | MassGather50 |
| **Use of Masks** |  |
| Protective mask use in public spaces/transport on voluntary basis (general recommendation not enforced) | MasksVoluntary |
| Protective mask use in public spaces/transport on mandatory basis (enforced by law) | MasksMandatory |
| **Other NPIs** |  |
| Teleworking recommendation or workplace closures | TeleworkingClosures |
| Closure of public spaces of any kind (including restaurants, entertainment venues, non-essential shops, partial or full closure of public transport, gyms and sport centers, etc). | ClosPubAny |

The mathematical model relies on contact data between the different age classes. For this, we obtained social contact matrices from Prem *et al* [17], which were derived from contact surveys and demographic data, and were available for all 32 European countries in our study. The age structure of each country has been retrieved from https://www.populationpyramid.net/. This data is released under a Creative Commons Attribution 3.0 (CC BY) license. Information on summer school holidays in each country was obtained from [18].

### Epidemiological COVID-19 transmission model

To model the evolution of the epidemic within each of the 32 European countries assessed, we built a model derived from a stochastic age-structured SEIR framework [12] considering 8 age classes (0-10, 11-20, 21-30, 31-40, 41-50, 61-60, and 70+ years). The population of each age class *i* is divided according to their infectious status: Susceptible (Si), Exposed (Ei, infected but not yet infectious), Detected infectious (Ii), Non-detected infectious (Ai) and recovered (Ri). Susceptible (Si) can become exposed upon age-specific force of infection λi:

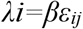

where *β* represent the transmission potential of each individual, which is modulated by the contact rate between age classes *ε*_*ij*_ as quantified by [17]. Then, after a latency period (1/ε) assumed to be 3 days, exposed individuals become detected infectious (Ii) according to probability *p* of case detection, which varies over time and has been estimated through a capture/recapture method ([19], see below). Otherwise, exposed individuals become non-detected infectious individuals (Ai). Finally, after an average of 5 days, individuals become recovered. Model simulations were performed using a τ-leap algorithm [20].

### Estimation of non-reported cases

We estimated daily non-reported cases for each country using the daily new reported cases and daily new deaths and the capture/recapture methodology described by Böhning and collaborators [19]. In brief, according to previous results in capture/recapture methodology [21], the following equality can be used:

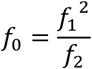

where *f*_*x*_ represents the number of individuals that have been observed *x* times. Translated into an epidemiological context, we are able to quantify how many cases at each time step *t* are not reported (*f*_*0*,_*H(t)*) based on the number of individuals notified only once (new cases detected, *f*_*1*_, Δ *N* (*t*)) and the number of individuals notified twice (new deaths reported, *f*_*2*_, Δ *N* (*t* − 1) − Δ *D* (*t*)):

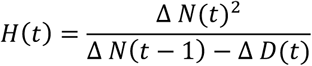

Using a bias-corrected form[21], we can use:

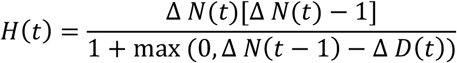

### Efficiency of PHRs at reducing COVID 19 transmission

We estimated transmission rates per country over time through Bayesian inference by running a particle Monte Carlo Markov Chain (pMCMC) with the epidemiological model described earlier. The chains had 10,000 iterations with a burn-in of 90% and a thinning of 1/10. The likelihood was computed with a particle filter of 100 particles and a negative binomial function of parameter 0.5 on the total daily number of new cases expected by the model and observed in the data (i.e., number of cases notified to the national public health system and number of undetected cases estimated by the capture/recapture method). The ratio between transmission rates with and without implementation of the PHR evaluated provides an estimation of the efficiency of this response, therefore assuming that it had an instantaneous effect.

### Efficiency of individual NPIs as part of larger PHRs

From the 258 PHR efficiencies estimated earlier, we first estimated in univariate linear regression analyses the effect on efficiency of having each NPI (presence/absence, dummy variables). Since the number of NPIs implemented was positively associated with response efficiency, we disaggregated these univariate analyses by the number of NPIs implemented as part of the response.

To isolate the efficiency of each NPI given the substantial overlap of NPIs implemented together, we transformed the NPI presence/absence and response efficiency dataset to reflect changes over time. For this, we first estimated for each country the difference in response efficiency with the previous time period (either no PHR or previous response) and the difference in NPI implementation (0 = no change; 1 = NPI added to the response; −1 = NPI removed from the response). We then estimated the effect of adding each NPI on the change in response efficiency using a multivariate linear mixed model that controlled for country’s GDP, number of interventions implemented, duration of implementation, the period of summer school holidays (fixed effects), and country of implementation (random intercept).

### Software and computing resources

All simulation and analyses were conducted using R 3.6 on the IRD itrop HPC (South Green Platform) at IRD Montpellier (https://bioinfo.ird.fr/). All code is available at https://github.com/Bisaloo/NpiEurope

## Results

Of the thirteen different NPIs implemented from February 1^st^ to September 16^th^, 2020 across the 32 European countries that reported data to the ECDC, bans on mass gatherings and school closures have been the most widely used, implemented by more than 90% of countries (figure 1A). These NPIs were fully implemented for a median time of three to five months. Across all the 258 PHRs analyzed, stay at home orders, bans on mass gatherings and school/university closures were typically implemented together (figure 1B). Despite large variability in terms of PHRs and sociocultural backgrounds across Europe, we were able to accurately quantify viral transmission rates over time in each country before and after implementation of PHRs, reproducing the observed national epidemiological dynamics (figure 2, correlations between observed and predicted cases were significant for every country, with an overall correlation coefficient of 0.58 and a p-value<2.e-16 across all countries). This allowed us to estimate the efficiency of the different country-level responses and isolate the effect of each NPI implemented in this period.

**Figure 1:**
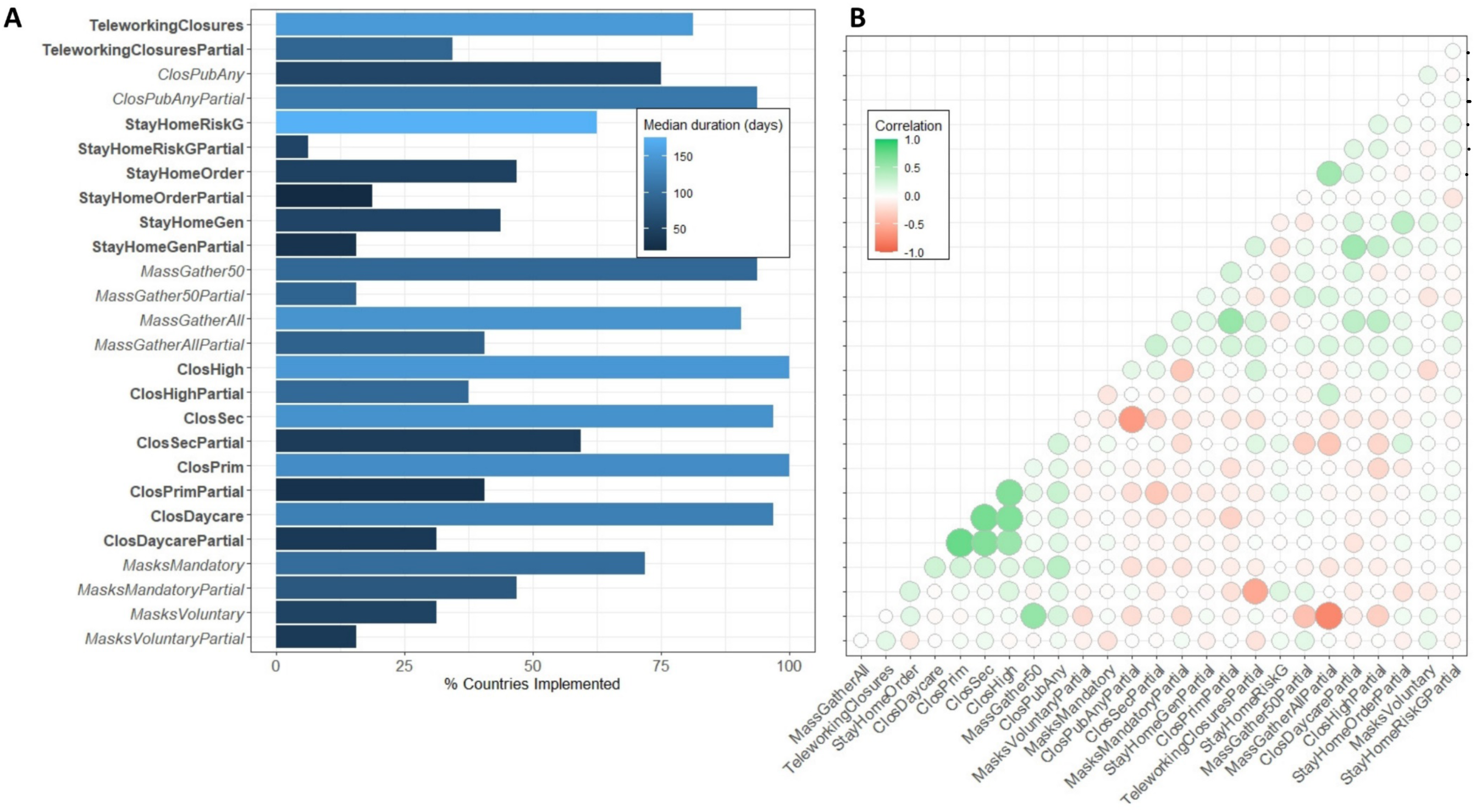
Implementation of NPIs against COVID-19 in Europe. (A) Status of implementation of 26 NPIs in 32 European countries between March and June 2020. (B) Correlation between implementation of each pair of NPIs as part of PHRs. A positive correlation (green circles) reflects that a pair of NPIs were commonly implemented together. A full description for each NPI is available in Table 1. Labels in X axis alternate between bold and italics to reflect different groups of NPIs.

**Figure 2:**
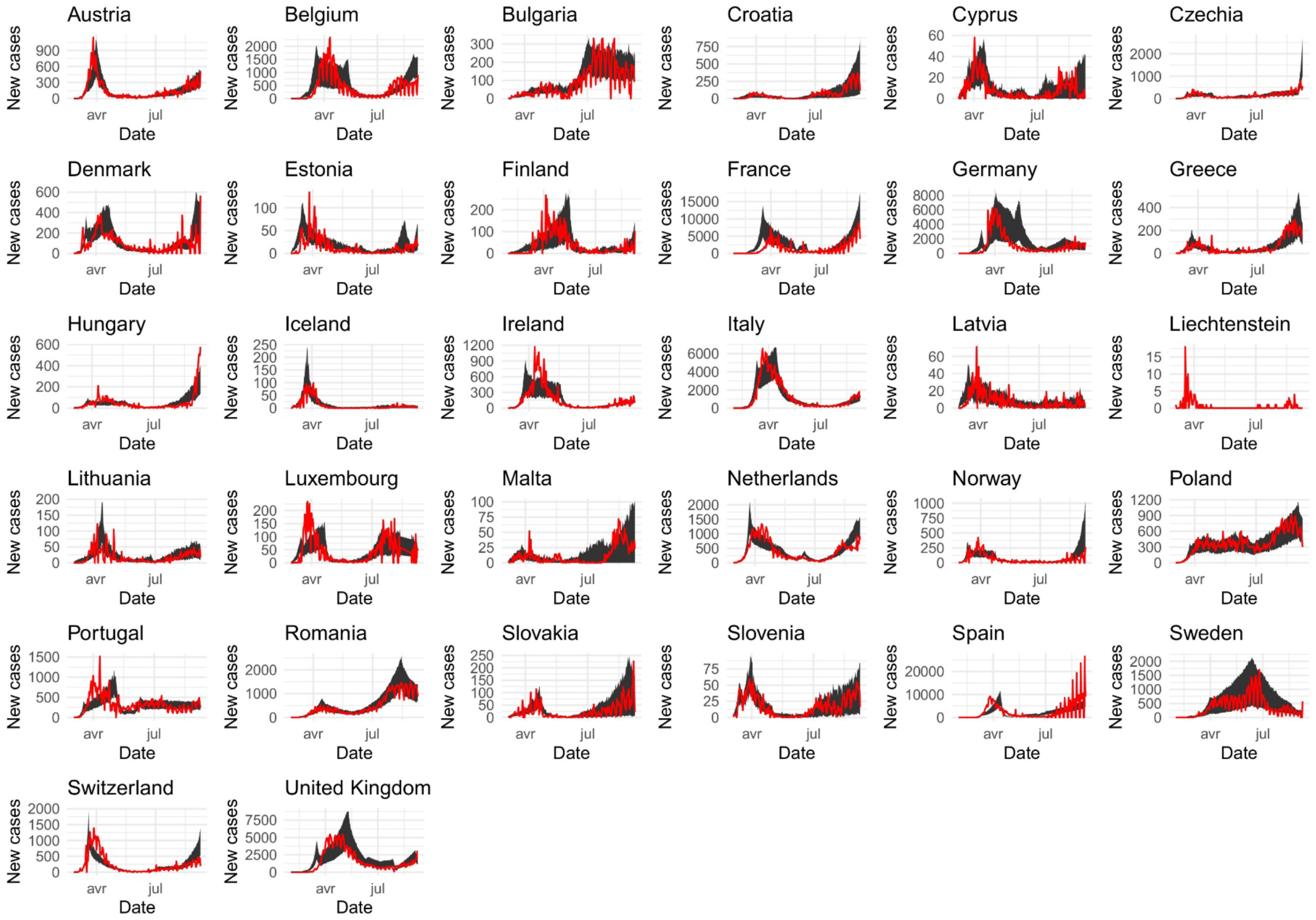
Model fit of COVID-19 incidence across European countries. Each panel shows model predictions (black shaded area) and confirmed number of new COVID-19 cases over time for each country, between February and September 2020. All estimations for each country are available in the section S1.

Overall, the combination of NPIs implemented as part of PHRs across Europe had a high efficiency at reducing transmission rates, with a median reduction of 71% and an interquartile range of 58-81%. With an estimated basic reproduction number *R0* of about 3, standard epidemiological models of COVID-19 suggest that to decrease the effective reproductive ratio below 1, transmission rates need to be reduced by at least 66% [22]. According to this, nearly two thirds of the PHRs evaluated could be part of a “suppression strategy” (figure 3A) [23], and many others could be part of mitigation efforts.

**Figure 3:**
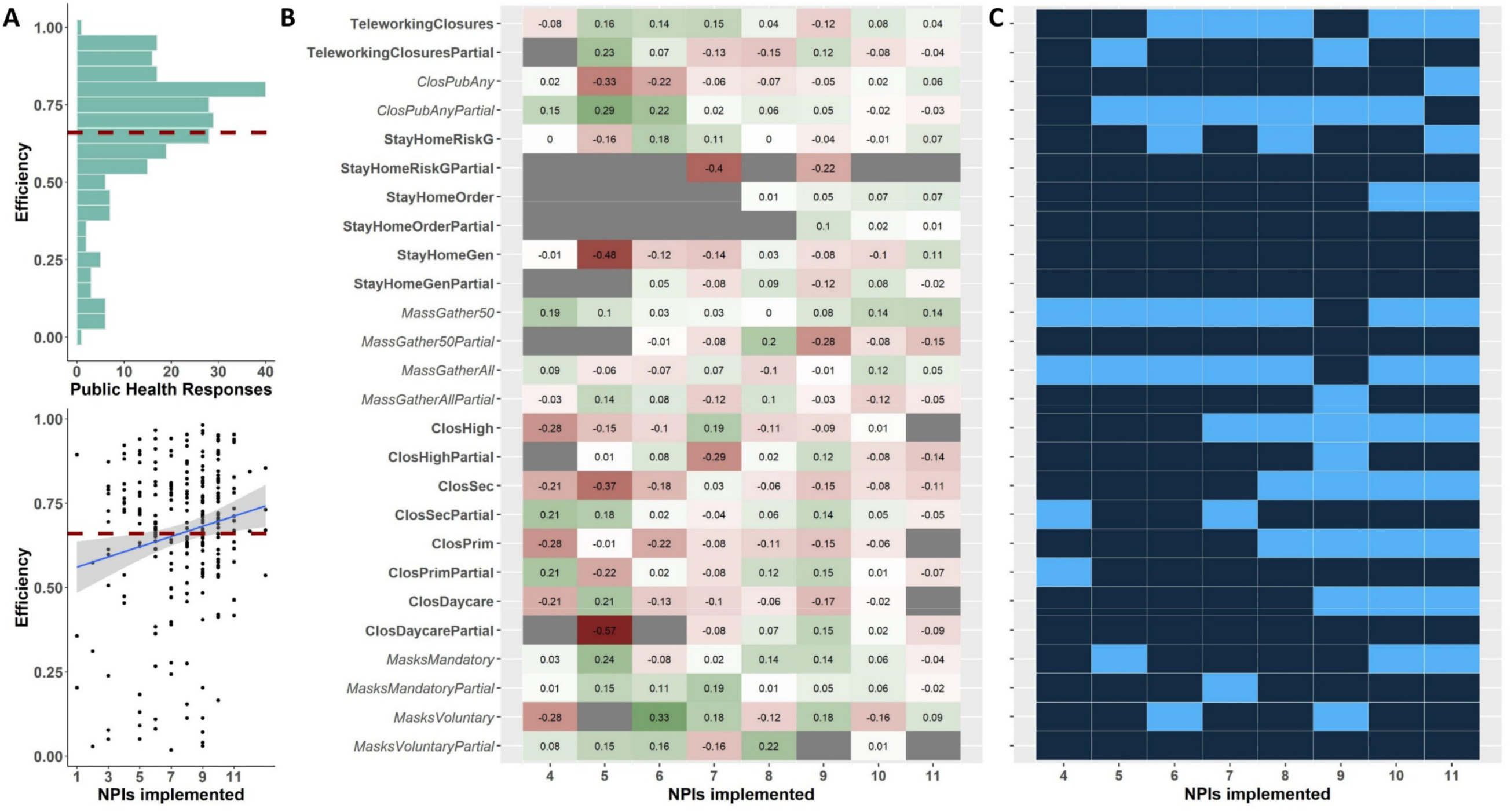
Efficiency of PHRs and NPIs implemented against COVID-19 in Europe. (A) Efficiency of PHRs, which has a positive correlation with the number of NPIs implemented simultaneously. Reductions in transmission above 66% (red dashed line) can achieve suppression given a R0 of 3. (B) Difference in response efficiency when a particular NPI was present (univariate models). (C) Shows which NPIs were part of the most efficient responses (light blue). Both B and C are disaggregated by the number NPIs that were implemented simultaneously. A full description for each NPI is available in Table 1. Labels in X axis alternate between bold and italics to reflect different groups of NPIs.

Efficiency was positively associated with the number of NPIs implemented as part of the PHR (figure 3A). To assess the effect of each NPI, we first estimated the added efficiency of each PHR when a particular NPI was present in univariate linear models, disaggregated by the number of NPIs implemented simultaneously (figure 3B). We found wide heterogeneity in efficiency between NPIs and for the same NPI at varying number of NPIs implemented simultaneously; only the presence of stay at home orders and strict bans on mass gatherings had consistent positive effects on PHR efficiency (figure 3B). In contrast, when we restricted the analysis to the most efficient PHRs, teleworking, bans to mass gatherings, and closure of businesses and services were the NPIs most consistently present across responses (figure 3C). Except for the closure of businesses and services, NPIs were seldom part of the most efficient PHRs when they had been partially relaxed.

Importantly, these efficiencies were estimated for countries with very different socio-cultural and economic backgrounds, NPIs were implemented for different lengths of time, and there was substantial overlap between them. To address this, we estimated the change in response efficiency over time when adding or removing a particular NPI, while controlling for GDP, number of NPIs, length of implementation, country of implementation and the summer holiday period (figure 4). We show that adding stay at home recommendations for risk groups to the PHR resulted in the largest increase in efficiency (β=0.24, 95%CI 0.16-0.32) followed by teleworking (β=0.23, 95%CI 0.15-0.31). Partial relaxation of these NPIs resulted in significantly lower than average response efficiency. In addition, enforced stay at home orders for the general population, bans to gatherings of over 50 people, and closures of non-essential businesses or services all resulted in significant increases in efficiency over 15%. Less constraining versions of these NPIs, such as limiting mass gatherings to less than 1000 people or non-enforced stay at home recommendations for the general populations led to smaller increases in PHR efficiency. Closure of universities and higher education establishments had a larger effect (β=0.23, 95%CI 0.06-0.22) than the closure of schools at secondary, primary or preschool levels. Recommendations for the use of protective masks, whether on a voluntary or mandatory basis, were the only NPIs that did not result in significant improvements in PHR efficiency. In contrast with teleworking and stay at home recommendations for risk groups, partial relaxation of all other NPIs resulted in non-significant changes in PHR efficiency.

**Figure 4:**
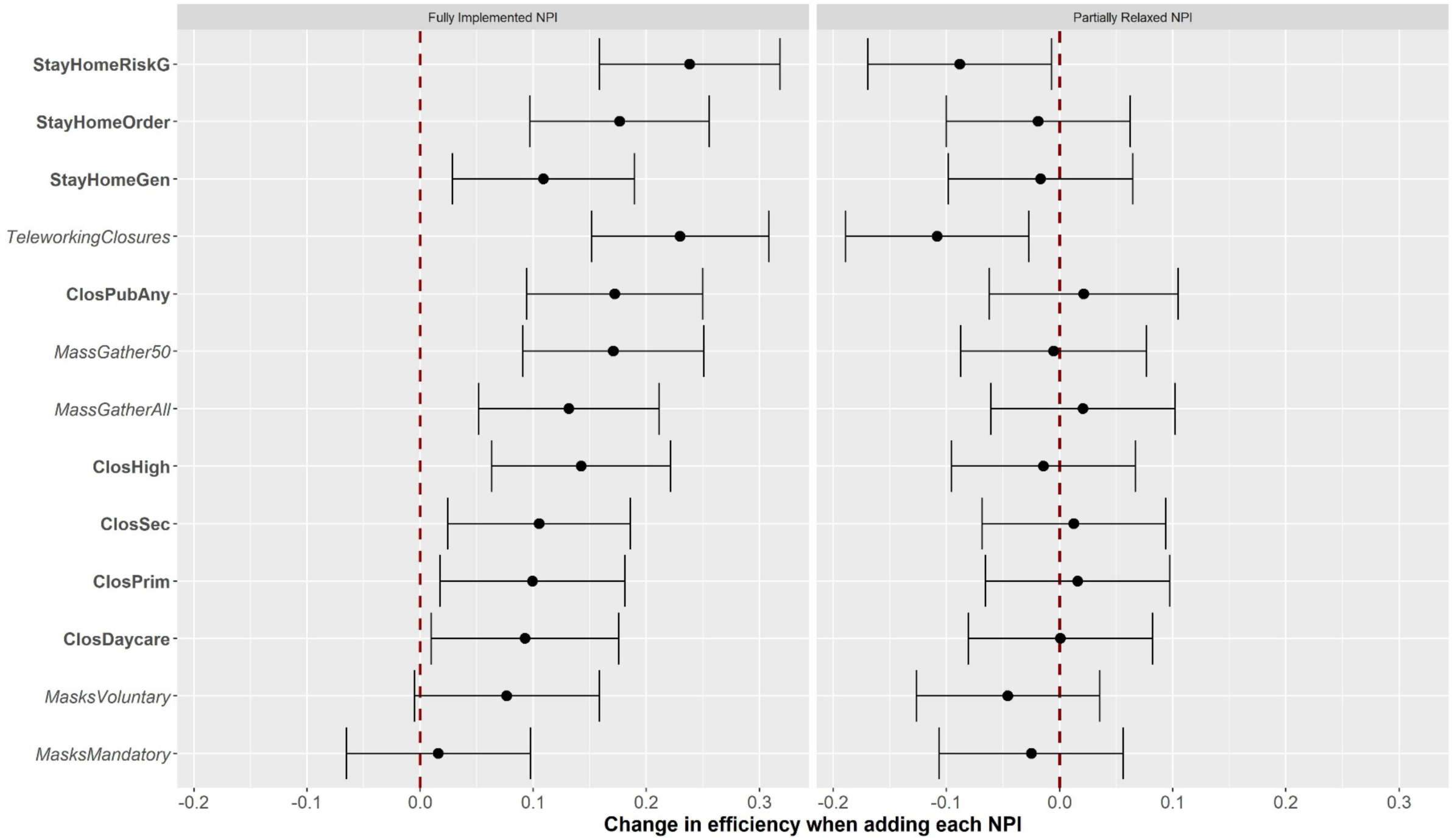
Additional PHR efficiency gained with NPIs implemented against COVID-19 in Europe. Results show the change in PHR efficiency over time when adding each of the 13 NPIs (mean adjusted effect and 95% confidence intervals in multivariate models). Results are disaggregated by level of implementation. A full description for each NPI is available in Table 1. Labels in X axis alternate between bold and italics to reflect different groups of NPIs.

## Discussion

Our empirical results on NPI implementation in 32 European countries show that most PHRs implemented between February and September 2020 had a sufficient efficiency to achieve suppression of the COVID-19 epidemic in the countries assessed. PHR efficiency increased with the number of NPIs implemented simultaneously, and the most efficient PHRs typically targeted multiple societal activities and population groups, including bans of gatherings, stay home orders, teleworking, and some form of school closure. This is consistent with previous modeling studies suggesting that packages of NPIs, not single interventions, are necessary to achieve epidemic suppression [5,7,13].

We find that enforced stay at home orders (i.e. lockdowns) significantly increased the efficiency of PHRs. However, nearly 90% of responses with efficiencies higher than 66% did not involve stay at home orders but a combination of other NPIs. In addition, adding stay at home orders to the PHR did not increase response efficiency more than other NPIs, such as bans to public gatherings, business closures, or teleworking. These results are in contrast with previous evidence indicating that the effect of lockdowns is substantially larger than other NPIs [4], and that only lockdown periods can reduce *R*_*eff*_ below 1 in order to suppress the epidemic [5]. This discrepancy can be explained by the fact that estimating empirically the relative efficiency of lockdowns compared to other combinations of NPIs requires large amounts of data, with a level of temporal and spatial heterogeneity in NPI implementation that has only recently been possible to achieve. In addition, integrated packages of NPIs that include combinations of school/university closures, teleworking, bans to gatherings and business closures can in practice reduce contact rates at the population-level in a way similar to lockdowns so that the additional efficiency gained by a lockdown, which is typically used as a last resort, is less pronounced in these contexts. Therefore, our study suggests that, although it is the most effective strategy at reducing COVID-19 transmission, complete lockdowns may not always be necessary to achieve epidemic suppression.

There has recently been increasing consensus around recommendations for the universal use of masks by the general population, as a cost-effective solution to allow activities in public places while minimizing the risk of viral transmission between individuals [24–26]. Although our analysis did not find that adding the use of masks to existing PHRs led to a significant increase in efficiency, the results should be interpreted with caution. Few European countries had implemented this type of NPI at the time of our analyses, and implementation was recent compared to other NPIs (figure 3). In addition, enforcement of the use of masks in Europe in the first months of the epidemic has been progressive with various level of intensity and, in contrast with many Asian countries, adoption of this individual protective measure by the population has been very variable. Contrary to the use of masks, we found that school closures at any level (daycare, primary, secondary) significantly increased PHR efficiency around 10%, although their effect was smaller than university closures and other NPIs targeting older age groups. There are still important knowledge gaps about the susceptibility of children to SARS-COV-2 and their capacity to transmit to others, with significant implications for decisions around reopening schools in the fall of 2020. Recent evidence from South Korea [27] and the United States [28,29] suggests that children may be as susceptible and transmissible as other age groups. Our results support this hypothesis, showing that closing schools to reduce contact rates among children and adolescents can be an effective way to increase the efficiency of PHRs at reducing transmission.

Data compiled by the ECDC shows that the heterogeneity in PHRs across countries was not only limited to the type NPIs implemented, but also their level of implementation. For instance, school closures or teleworking have been partially relaxed by including daily rotation in physical attendance in order to reduce the number of people present at facilities each day. Similarly, stay at home orders have been partially relaxed by slotting particular times of the day for different age groups to go out. Yet, previous studies have failed to consider this when measuring the efficiency of NPIs [5,7,13]. Our results suggest that implementation of partially relaxed NPIs can either have null added value to national PHRs or even result in a reduction of PHR efficiency (e.g. teleworking measures and stay at home recommendations for risk groups). This could have important implications for future national responses, as NPIs implemented in Europe during the study period were partially relaxed during nearly one fourth of the time.

This study had several limitations. First, we used an age-structured model where each age class has the same susceptibility to the virus, and we assume that detected and non-detected cases have the same transmissibility capacity because non-detected cases can be both asymptomatic and symptomatic cases. However, it is still unclear whether asymptomatic cases and children have similar or lower SARS-COV-2 transmissibility capacity compared to other population groups, which could affect our results. Second, although our mathematical model allowed us to accurately quantify PHR efficiency, responses were comprised of 13 different NPIs that frequently overlapped over space and time. We estimated the added efficiency provided by each NPI via statistical time-series analyses controlling for relevant factors, but these should not be interpreted as NPI independent efficiencies because we could not control for the implementation of other NPIs in our multivariate framework. Given the substantial overlap in NPI implementation, this could also lead to biases in the estimated efficiency of certain NPIs, which could explain for instance the small effect of lockdowns as compared with stay at home recommendations for at-risk groups. Third, we use a comprehensive database on NPI implementation compiled by the ECDC from country reports, but other interventions such as travel restrictions could have impacted epidemic progression at European level [8,30]. This could have biased the estimates for the NPIs evaluated here.

During subsequent epidemic waves, our study suggests that comprehensive PHRs that include stay at home recommendations for risk groups, teleworking, bans to gatherings of 50 people or more, closures of businesses, services and universities will be most effective at suppressing the epidemic. Stay at home orders substantially contribute to response efficiency but may not be necessary to achieving epidemic suppression. Further evidence is needed to assess whether enforcing the use of masks significantly improves overall PHR efficiency at the country level.

## Data Availability

Data are available upon request

## Acknowledgments

HG and BR are supported by a grant from the “Agence Nationale de la Recherche” (ANR-DigEpi). We thank Simon Cauchemez for insightful comments on earlier versions of this manuscript.

## Notes

### Competing Interest Statement

The authors have declared no competing interest.

### Funding Statement

This study has been supported through a grant from the Agence Nationale de la Recherche (ANR-DIGEPI)

